# Single-workflow Nanopore whole genome sequencing with adaptive sampling for accelerated and comprehensive pediatric cancer profiling

**DOI:** 10.1101/2025.10.02.25336569

**Authors:** Nicholas Geoffrion, Charlene Lawruk-Desjardins, Sylvie Langlois, Marjorie Aleman Alvarado, Niklas Dreyer, Anthony Claude Carrier, Véronique Lisi, Chantal Richer, Alex Richard St-Hilaire, Pascal Tremblay-Dauphinais, Alain R. Bataille, Thomas Sontag, Séverine Landais, Alexandre Rouette, Loubna Jouan, Imène Boumela, Banafsheh Khakipoor, Sandy Fong, Stéphanie Vairy, Catherine Goudie, Nada Jabado, Raoul Santiago, Adam Shlien, Martin A. Smith, Daniel Sinnett, Sonia Cellot, Thai Hoa Tran, Vincent-Philippe Lavallée

## Abstract

Timely and comprehensive molecular classification is critical for therapeutic decisions in pediatric oncology. However, current diagnostic workflows rely on multi-step testing and are resource- and time-intensive. We present whole-genome sequencing with adaptive sampling (AS-WGS) protocol using Oxford Nanopore Technologies optimized for pediatric oncology, enabling unified detection of genomic, structural, and epigenomic alterations in a single assay. Applied to 31 pediatric cancer patient samples, AS-WGS achieved high on-target coverage across hundreds of loci of interest for identifying somatic anomalies, while maintaining pan-genomic coverage for copy number assessment and methylome data. We demonstrate that AS-WGS, as a single approach, captures all categories of clinically relevant alterations, including most copy number changes, fusions, and mutations, even subclonal ones. Time stamp analyses revealed that clonal alterations are confidently supported within the first sequencing day, sometimes within the first hours. We developed and reported an open-source bioinformatic pipeline (nf-core-oncoseq) that facilitates streamlined and fully integrated analysis. This approach consolidates complex testing into a single, rapid assay, enabling near real-time cancer characterization. Our findings support AS-WGS as a transformative diagnostic platform for pediatric oncology.

## INTRODUCTION

Pediatric cancers, including solid tumors and leukemias, are driven by a broad spectrum of genomic and epigenomic alterations, such as single nucleotide variants (SNVs), gene fusions, copy-number variants (CNVs), intragenic deletions and amplifications, and DNA methylation changes^1–3^. Accurate and timely characterization of these alterations is essential for establishing the precise diagnosis, informing risk stratification and guiding correct therapeutic decisions^4^. Current diagnostic workflows rely on multi-step iterative testing, including cytogenetics and molecular-based approaches, each providing distinct but complementary information. While some targeted assays yield rapid results, others (e.g., whole exome sequencing or WES) typically require more than 2 weeks to obtain data, potentially delaying risk assessment and important treatment decisions. This disjoint approach also increases costs, consumes limited tumor material, and complicates data integration, highlighting the need for unified, rapid, and cost-effective strategies. Whole Genome Sequencing (WGS) is increasingly recognized as a more comprehensive diagnostic strategy^5–8^. However, its implementation in clinical laboratories also remains challenging due to the need to deliver results within clinically relevant timeframes and the relatively lower sequencing depth (∼80X) compared to other sequencing approaches.

Long-read sequencing using Oxford Nanopore Technologies (ONT) offers a promising alternative to the current approaches^9^. ONT provides direct sequencing of native DNA, enabling structural variants detection^10^ and haplotype phasing, while preserving methylation marks^11^. ONT data can be analyzed simultaneously during sequencing, as soon as the data is generated, holding great promise for faster identification of genomic alterations^9, 12^. WGS with adaptive sampling mode (AS-WGS) enables selective enrichment of predefined genomic regions, improving depth of coverage for target loci of interest while retaining genome-wide context^13, 14^. Prior efforts have demonstrated promising results for structural variants detection using AS-WGS in the context of digital karyotyping^12^, but the performance of this strategy did not support small variant identification, an important feature in oncology. Furthermore, while methylation-based classifiers have shown potential for early diagnoses in brain and hematologic malignancies, their performance under adaptive sampling conditions remains untested^15–18^. Finally, existing bioinformatics pipelines are not designed to support integrated somatic variant calling, structural variant detection, and methylation-based classification, limiting their ability to fully leverage the real-time potential of adaptive sampling data.

Here, we report an AS-WGS protocol tailored for pediatric oncology, which was evaluated in a standardized way on 31 consecutive biological samples against extensive clinical testing. We developed an open-source workflow, nf-core-oncoseq^19^, to enable integrated and real-time analysis of somatic mutations, structural alterations, copy number variants, and methylation patterns from AS-WGS data. Our results demonstrate the feasibility of using a single AS-WGS-based assay to achieve rapid and complete identification of every type of alteration supporting the molecular characterization of pediatric cancers, with important implications for diagnostic efficiency and clinical decision-making.

## METHODS

### Cohort and clinical testing

Patients were enrolled in the SIGNATURE study, initiated in November 2019, a provincial childhood cancer precision medicine program enrolling patients from all four pediatric oncology centers in Quebec, Canada. All patients underwent conventional clinical testing as per institutional standards including Fluorescence in situ hybridization (FISH), G-banding karyotype, Comparative Genomic Hybridization (CGH), Reverse Transcription (RT)-PCR, WES and/or total RNA sequencing. Additional details for cohort and clinical genomics workflow^20, 21^ are presented in Supplementary Methods and Table S1.

### ONT Adaptive sampling

#### Panel Preparation

Target panels were constructed following the recommendations from ONT^22^. We selected 357 genes relevant to pediatric oncology^23^ and added up to 23 genes to better cover the landscape of alterations in acute myeloid leukemias (AML). To ensure adequate coverage around each target, we included a 20 kb buffer region upstream and downstream, which aligns with the library’s target N50 (see sample preparation, below). To facilitate the rapid addition of genes or loci and the buffer region to the panel, we developed the QuickPanel.R script, available (https://github.com/chusj-pigu/gene-panels). Altogether, the panels used in this study encompass 375 to 380 loci on the GRCh38 human reference genome representing approximately 1.7% of the human genome (Table S2), in accordance with the optimization guidelines of ONT^22^. The size of the panel tolerates the addition of genes while remaining within the proportion recommended by the manufacturer.

#### Sample preparation and sequencing

DNA extraction, shearing to ∼12 kb fragments using Covaris G-tubes, quantification and library preparations are detailed in Supplementary Methods. Each sample was loaded onto a single FLO-PRO114M flow cell, sequenced on the PromethION 24 at the CRA-CHUSJ Integrated Genomics Meta-Platform (MPGI) and data was processed using standard workflows^24, 25^. Additional details, including flow cell wash and reload procedures, are provided in the Supplementary Methods.

### Bioinformatics

We developed a real-time bioinformatics workflow (nf-core-oncoseq) optimized for AS-WGS, enabling timepoint-specific variant detection (Figure S1, https://github.com/chusj-pigu/nf-core-oncoseq). The workflow, based on existing tools^25–39^, supports analysis at different sequencing intervals and includes CNV, structural variants, and SNV/indels calling. Additionally, we created VarClock^40^, a tool for timestamped read-level variant tracking, enhancing mutation detection across timepoints. Additional bioinformatic details are provided in Supplementary Methods.

## RESULTS

### AS-WGS enables robust enrichment of clinically relevant target regions for pediatric oncology

A total of 31 samples from 30 pediatric patients were included. Of those, 27 represented tumor samples from 20 hematological malignancies (B-cell acute lymphoblastic leukemia (B-ALL) [n=8], T-ALL [n=3], AML [n=7], Burkitt leukemia [n=1] and myelodysplastic syndrome [n=1]) and seven solid tumors (Table S1). Four additional samples consisted of normal bone marrow, used for quality control assessment. Following uniform library preparation, all samples underwent AS-WGS for 72 hours, with selective enrichment of 375 to 380 genes or loci followed by nf-core-oncoseq data processing (Table S2). Mean coverage achieved was 165X for target autosomal regions (range 58-256X), and 18X in pan-autosomal genomic regions (outside of the target regions, range 10-26X) (Figure 1A). Including 20-kb buffer regions upstream and downstream of targets allowed for stable coverage across gene exons, while also capturing introns and adjacent regulatory regions (Figure 1B).

**Figure 1.**
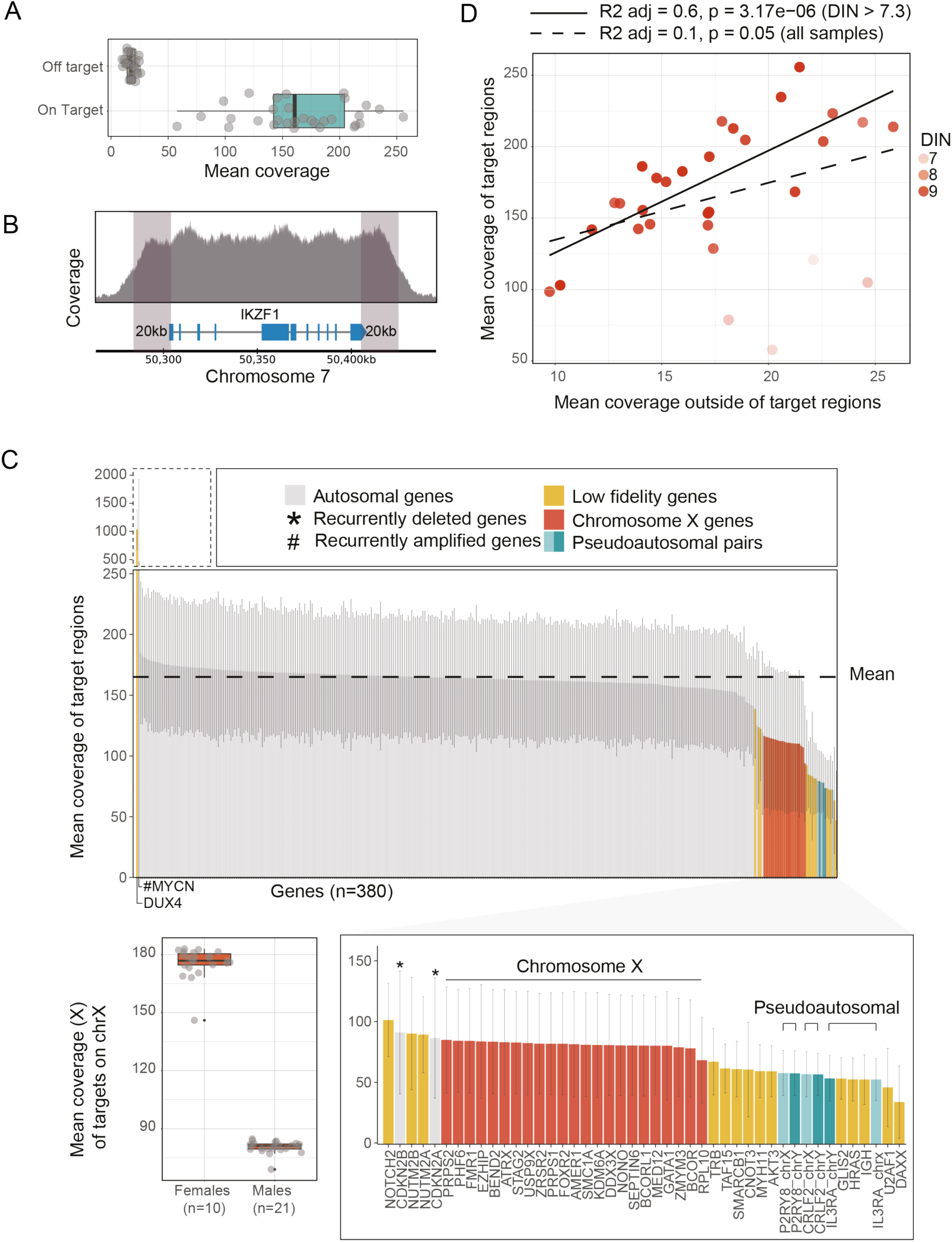
Adaptive sampling consistently improves coverage across target regions. **A)** Boxplot of mean coverage achieved for each sample (n=31) inside targeted region ("On Target”) and outside targeted region ("Off target”). **B)** Gene coverage across a typical target region. Coverage changes in the 20kb buffer zones, highlighted in gray, reaching maximum depth over the genic region (shown in blue under the coverage graph). **C)** Mean coverage across samples (n=31) for each gene/locus in targeted panel, ordered by decreasing value. Vertical error bars show the standard deviation, and colors highlight known outliers (low fidelity genes) and genes on sexual chromosomes, expanded underneath. Boxplot of mean coverage of the X chromosome split according to sex. **D)** Mean coverage on target region as a function of mean coverage off target (n = 31), opacity shows gDNA DIN of each sample.

Across the entire cohort, target coverage was uniform for 333 out of 380 (88%) genes (Figure 1C, Table S3). Of the remaining genes, 23 were located on the X chromosome, which had the expected reduced coverage in males (red bars and boxplot in Figure 1B). This reduced coverage is not expected to impact variant identification, as these genes will be hemizygous. Additionally, uniformity of 3 genes (*CDKN2A*/*B* and *MYCN)* was impacted by recurrent deletions and amplifications, respectively, while it remained stable in copy neutral samples (Figure 1C and Figure S2). The remaining 21 genes (5.5% of targets) were genes whose coverage originated mostly (>50%) from non-uniquely mapped reads and were classified as ‘low fidelity’ (Figure 1C and Table S2). These genes were predominantly pseudoautosomal pairs (n=3) and were enriched in genes with pseudogenes or highly variable regions. For the 336 high fidelity autosomal genes, coverage of target regions was highly uniform, with on average 98% of genes in each sample falling within a z-score of < 2.75 in normal control samples (Figure S3). This further emphasizes the stability of coverage across individual samples.

On-target and pan-genomic coverage were highly concordant, highlighting inter-experiment variability in coverage (Figure 1D), and we therefore sought to identify the factors underlying this variability. As anticipated, DNA quality, particularly samples with a DNA integrity number (DIN) below 8, consistently resulted in reduced target coverage (Figure S4A). Notably, a positive correlation was observed between the number of active nanopores and final target coverage (Figure S4B). In addition, flow cell reception date, which reflects flow cell batch, was also independently associated with variability in coverage, regardless of DNA quality or pore count (Figure S4C). This finding was further supported by repeated sequencing of a sample with decreased coverage in a second experiment, using the same protocol (red dot in Figure S4C). No other factors were identified as influencing sequencing coverage outputs (detailed experimental provided in Table S4).

Despite inter-sample variability, our protocol achieved unprecedented overall stable target enrichment, including intronic and upstream/downstream regions.

### AS-WGS single-test identifies most clinically relevant alterations in pediatric oncology

We next evaluated the performance of AS-WGS in detecting alterations across multiple variant classes, using as reference the findings from conventional diagnostic approaches (Figure 2A and Table S5).

**Figure 2.**
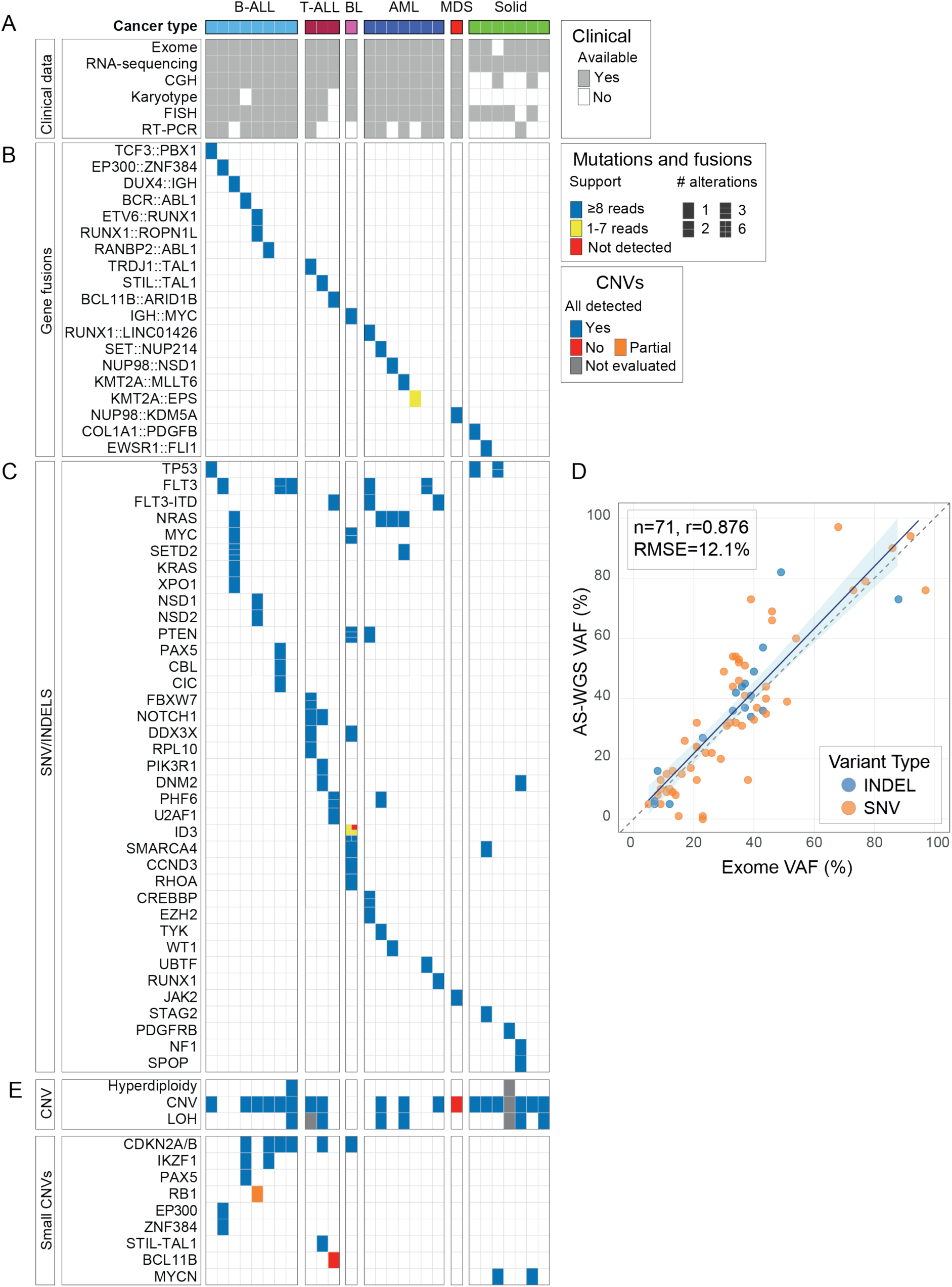
Performance of AS-WGS in comparison to clinical genomics. Alteration map across cancer types for all 27 patient samples. Each column represents one patient. **A)** Clinical tests performed for each sample. Panels B, C, and E display results from clinical testing for gene fusions (B), mutations (C) and copy number variations (E). Detection quality by AS-WGS is color coded: blue cells indicate events supported by at least 8 alternative reads for fusions and mutations and confidently called CNVs based on visual inspection. **D)** Correlation between variant allele frequencies (VAFs) identified by WES compared with AS-WGS for single nucleotide variants (SNVs) and insertions/deletions (indels). Cancer types are abbreviated as follows: ALL (acute lymphoblastic leukemia), AML (acute myeloid leukemia), BL (Burkitt leukemia), MDS (myelodysplastic syndrome). LOH (loss of heterozygosity).

#### Gene Fusions

Gene fusions are defining hallmarks of oncology and therefore represent a class of alterations that AS-WGS must detect. Globally, out of the 19 gene fusions reported in the clinical data (Figure 2B), 18 (95%) were confidently supported by AS-WGS, with a mean of 56 reads (range: 24–112). The only undetected fusion, *KMT2A*::*EPS15*, was supported by a single read and occurred in a specimen with low cellularity and DNA integrity (DIN = 6.9). Most AS-WGS supported fusions (16/18) were confidently called by the structural variant detection algorithm (Sniffles2^31^), while the remaining two were identified upon visual inspection of raw data (Figure S5A), highlighting areas for improvement in algorithmic detection of fusions. As previously reported^12^ *DUX4*::*IGH* rearrangements posed specific mapping challenges due to low-confidence alignments at both loci, highlighting a need for systematic querying of such events in pipelines.

#### Single-Nucleotide Variants and Indels

Across the cohort, 72 SNVs and small indels with a variant allele frequency (VAF) ≥ 5% were reported by clinical WES testing (Figure 2C). Of those, 68 (94%) were confidently detected by AS-WGS (≥ 8 supporting reads), with an average read depth of 156 reads [range 39–273]. The remaining 4 variants were all located in *ID3* within a single Burkitt leukemia sample: three were weakly detected (1–7 reads), and one was not detected. This likely reflects variable subclonality in different patient-derived specimens as this region was well covered by AS-WGS (265X). No additional discordances with clinical data were observed, and thus all reported mutations were successfully identified in 21 of 22 patients (95%) harboring SNV/indels. Importantly, VAFs were highly concordant between AS-WGS and WES, confirming that clonal and subclonal architectures were faithfully preserved (Figure 2D). AS-WGS also detected technically challenging indels, including internal tandem duplications (ITDs) in *FLT3* (n=3) and *UBTF* (n=1). While most mutations were identified through nf-core-oncoseq variant-calling pipeline (57/72, 80.5%), others (n=8) were detected by DeepVariant^36^, underscoring the value of multi-caller integration. The remaining 6 variants had low read support (2-11) but could be visualized in the read data via IGV (Figure S5B). Altogether, these results demonstrate that optimization of the AS-WGS protocol directly enhances coverage and thus sensitivity for both clonal and subclonal variants, several of which carry diagnostic or prognostic significance in pediatric cancers.

#### Copy Number Alterations

Every reported large CNVs was accurately detected in 16 of 17 samples, including recurrent and prognostically relevant alterations such as hyperdiploidy (Figure 2E). The missed CNV consisted of a subclonal (∼ 25% according to microarray) interstitial chromosome 12 deletion. AS-WGS showed strong performance in identifying focal deletions, with reliable detection of gene-level or intragenic deletions in 14 out of 16 samples, including all *CDKN2A*/*B* (n=6), *PAX5* (n=1) and *IKZF1* (n=2) alterations, which are recurrent and/or prognostically relevant in ALL. High-level amplifications, such as *MYCN* in neuroblastoma, were also robustly detected. Of the two missed small-sized alterations one was an intragenic duplication of *BCL11B*, of unknown clinical significance. This event occurred in association with a Tier 1 *BCL11B*::*ARID1B* rearrangement, which was adequately supported and identified. The other missed alteration was in a sample harboring an intragenic *RB1* deletion in addition to a gene-level *RB1* deletion, the latter being the only alteration detected by AS-WGS.

Taken together, AS-WGS demonstrated high sensitivity in detecting somatic alterations across every major variant class, including SNVs/indels, gene fusions, and copy-number changes. The assay, based on complete (72h) sequencing data, reliably identified alterations at VAFs as low as 5% and captured complex structural events and focal deletions, establishing it as a powerful single-test alternative to the current multi-platform diagnostic workflow.

### Identification of most alterations in less than 24 hours using adapted workflows

Optimization of the nf-core-oncoseq workflow enabled the results described above to be generated in under five days, with the following breakdown: ∼3 hours for DNA extraction, ∼3 hours for library preparation, 72 hours for sequencing, and ∼24 hours for nf-core-oncoseq processing and data analysis. As ONT sequencing allows real-time data acquisition, we retrospectively assessed the time required to detect various types of alterations (Figure 3). Using timestamp information embedded in the raw data, we found that oncogenic fusions within panel genes could be identified with at least 5 reads support between 18 minutes and 21 hours (Figure 3A). Most (17/18, 94%) fusions reached ≥10 supporting reads in less than 24 hours. Similarly, SNVs and indels supported by a minimum of 5 alternate reads were detected between 36 minutes and 52 hours. As expected, the time to confident detection was influenced by VAF, with clonal mutations (VAF >25%) showing a narrower detection time window ranging from 36 minutes to 19.5 hours, with a 5-read average time of detection of 1.79 hour (Figure 3B).

**Figure 3.**
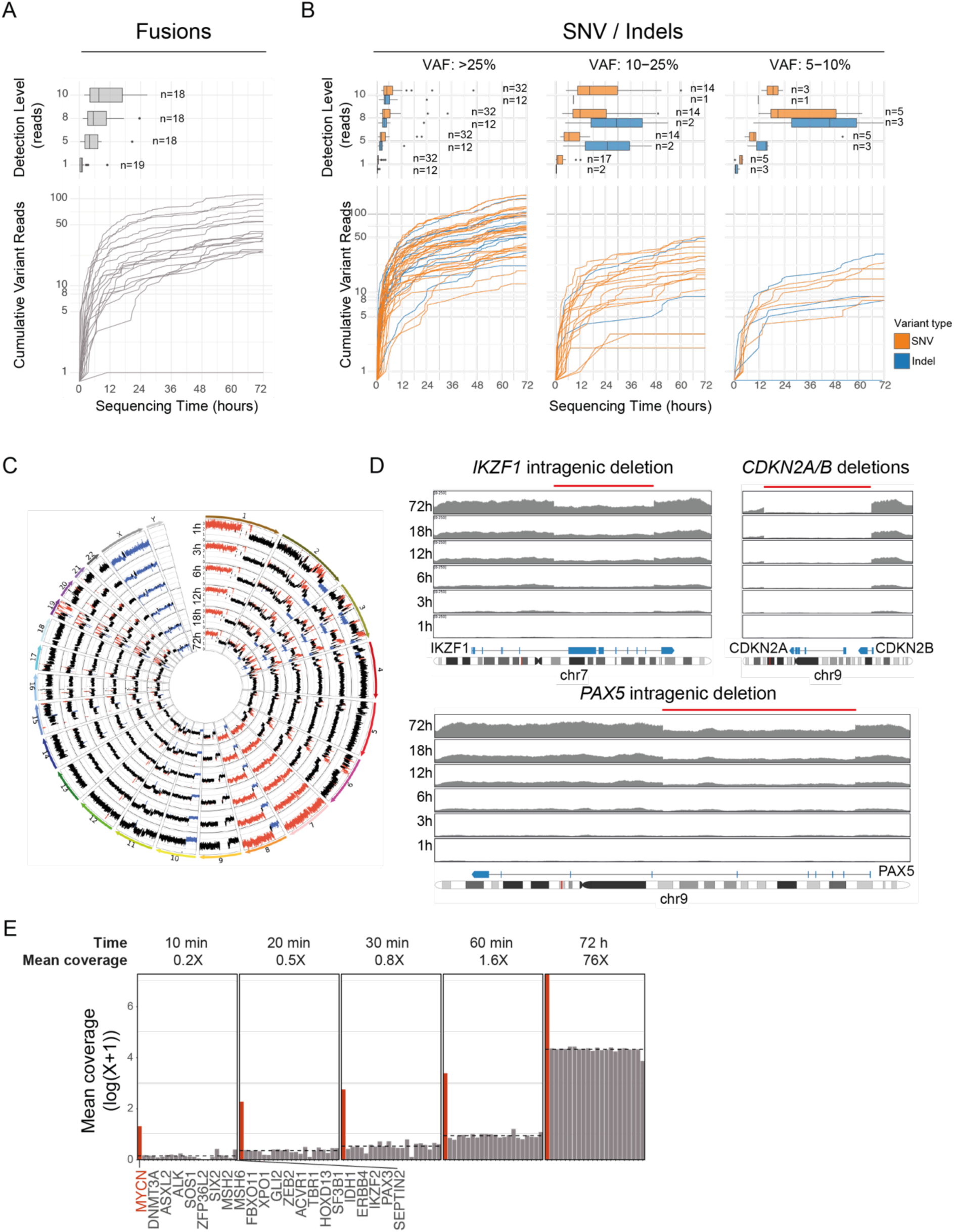
Alteration supports over sequencing time. **A)** Mean time required to reach predefined depth threshold for fusions (top) and fusion supporting read depth over time (bottom). **B)** Mean time required to reach predefined depth threshold for SNV and indels based on clonality (clonal VAF >25%, subclonal VAF 10–25%, highly subclonal VAF 5–10%) (top) and variant supporting read depth over time for each sample. **C)** Representative circos plots at selected time points (time decreasing from middle) highlighting early stable detection of large CNVs (osteosarcoma sample SI_ASWGS0534; del: blue, gain: red). **D)** Representative example of coverage at selected time points of on target regions showing early detection of intragenic deletions of *IKZF1* and *PAX5* and gene level deletion of *CDKN2A* and *CDKN2B* (B-ALL sample SI_ASWGS0318). **E)** Representative example of coverage of chromosome 2 genes showing early identification of increased *MYCN* coverage, reflecting gene amplification (neuroblastoma sample SI_ASWGS0705). Red represents a coverage with a Z-score > 2.75. For each timepoint, mean coverage of *MYCN* was 3, 9, 15, 29 and 1451 respectively. All subpanels display the same gene order across timepoints.

For CNV detection, we analyzed time-based downsampled data at the intervals indicated in Figure 3C-E. Large CNVs were all consistently identifiable in under one hour of sequencing (Figure 3C), supporting the feasibility of same-day reporting. Monogenic and intragenic deletions were also reliably detected, typically within 3-12 hours of sequencing (Figure 3D). Gene amplifications were suspected in the first 10 minutes of sequencing, with confident signals after 20-30 minutes (Figure 3E). Altogether, these results highlight that most clinically meaningful results are present in the data within the first day, sometimes in the first hours, and that nf-core-oncoseq workflow can capture these alterations while sequencing is ongoing.

### AS-WGS supports methylation clustering

The PCR-free approach used for AS-WGS preserves native methylation, thereby enabling the observation of base modifications such as 5-methylcytosine and 5-hydroxymethylcytosine. Such modifications, typically assessed by methylation arrays, have been used to classify solid tumors in real time^15, 16^. Recently methylation-based classifiers have been developed for and tested on ONT WGS datasets, including in leukemia (MARLIN^17^ and ALMA^18^). These tools have not previously been tested on AS-WGS datasets.

MARLIN and ALMA (v0.1.14) were applied to our AS-WGS data and were each able to accurately predict the leukemia’s lineage in over 70% of samples (ALMA:15, MARLIN: 14 of the 19 samples) (Figure 4A and Table S6). Overall, for 17 of the 19 samples, the lineage was properly identified by at least one of the two classifiers. ALMA predicted the correct lineage in 7 of the 8 AML classes. MARLIN performed particularly well on the ALL samples, identifying the correct lineage for all but one T-ALL sample (10/11) misclassified as acute leukemia of ambiguous lineage. Of relevance, the T-ALL sample was an Early T-cell Precursor-ALL case expressing both T-cell and myeloid antigens, which could lead to the lineage misclassification by MARLIN.

**Figure 4.**
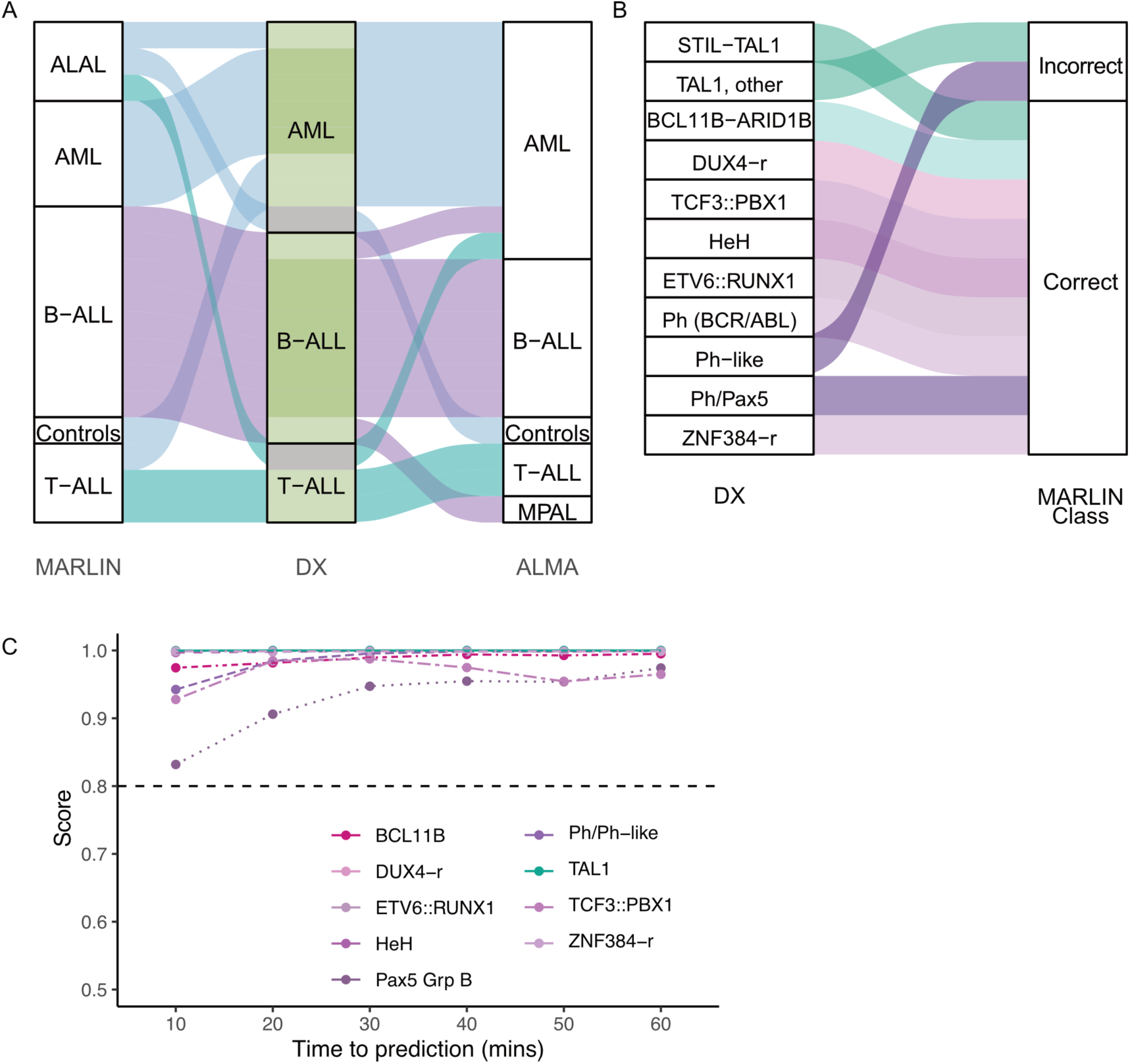
Methylation-enabled rapid classification of pediatric leukemias. **A)** Comparison of clinical data based diagnostic lineage (center) and methylation-based lineage classification using MARLIN (left) and ALMA (right). Alluvium is filled based on true diagnostic lineage. Diagnostic lineage is fill according to the number of correct predictions. **B)** Comparison of clinical subgroup and MARLIN methylation class. Alluvium is colored based on clinical subgroup. **C)** Prediction score of MARLIN over time for the correct predictions for each of the ALL samples correctly categorized by MARLIN (n=9). Data is color coded according to MARLIN methylation class.

We next evaluated the capacity of classifiers to accurately assign samples to their respective molecular subgroups, several of which carry important diagnostic and prognostic significance. This was only possible for ALL, as our AML cohort largely comprise non-recurrent cytogenetic abnormalities for which labels are not included in published classifiers. In B-ALL, MARLIN perfectly identified the family in all eight samples, and the class in seven of the eight samples (Figure 4B). Only one Philadelphia (Ph)-like sample was misclassified as *PAX5*-r, while all the following subgroups were all adequately predicted: hyperdiploidy, *ETV6*::*RUNX1*, *DUX4*-r, *ZNF384*-r, *TCF3*::*PBX1*, Philadelphia (Ph)/Ph-like, *PAX5* altered. In T-ALL, MARLIN predicted family or class in two of the three cases (*STIL*::*TAL1*, and *BCL11B*-r), while mislabeling *TAL1*-r into *TLX1*-r subgroup.

To evaluate the speed of methylation-based classification and its relevance to clinical decision, we performed a timestamp-based down sampling of the BAM files for each 10-minute period in the first hour of sequencing. This experiment revealed that all methylation-based predictions were already accurate after only 10 minutes of sequencing (Figure 4C). These results highlight that AS-WGS data supports methylation classifiers and, consequently, enables rapid and multi-modal diagnostics: classification predictions can be generated in the first sequencing hour, and over the course of the remaining sequencing duration, subtype-defining alterations can be rapidly corroborated and validated.

### Sequential dynamic analysis of AS-WGS data

Our results inform the optimal sequence of analyses: methylation classifiers and CNV detection can be performed within the first hour of sequencing, clonal variants and fusions are detectable within the first 24 hours, while subclonal variants require the full 72-hour sequencing period for reliable detection (Figure 5A). This chronology is illustrated through two representative samples. In the first, a leukemia sample was adequately classified as T-ALL with *TAL1* rearrangement within 10 sequencing minutes, with CNV profile stabilizing at 1 hour. *STIL*::*TAL1* fusion reads were supported at 3 hours, and clonal SNVs, including *NOTCH1*, were detected in under 6 hours. The 72-hour pipeline further evidenced CN-LOH (Figure 5B). In the second sample, a solid tumor, the *EWSR1*::*FLI1* fusion reached confident read support at 6 hours, alongside clonal somatic mutations in *STAG2* and *SMARCA4*. A subclonal *TP53* mutation required the complete 72-hour dataset to reach confident support (Figure 5C). These findings highlight the applicability of dynamic AS-WGS analysis to both solid tumors and blood cancers.

**Figure 5.**
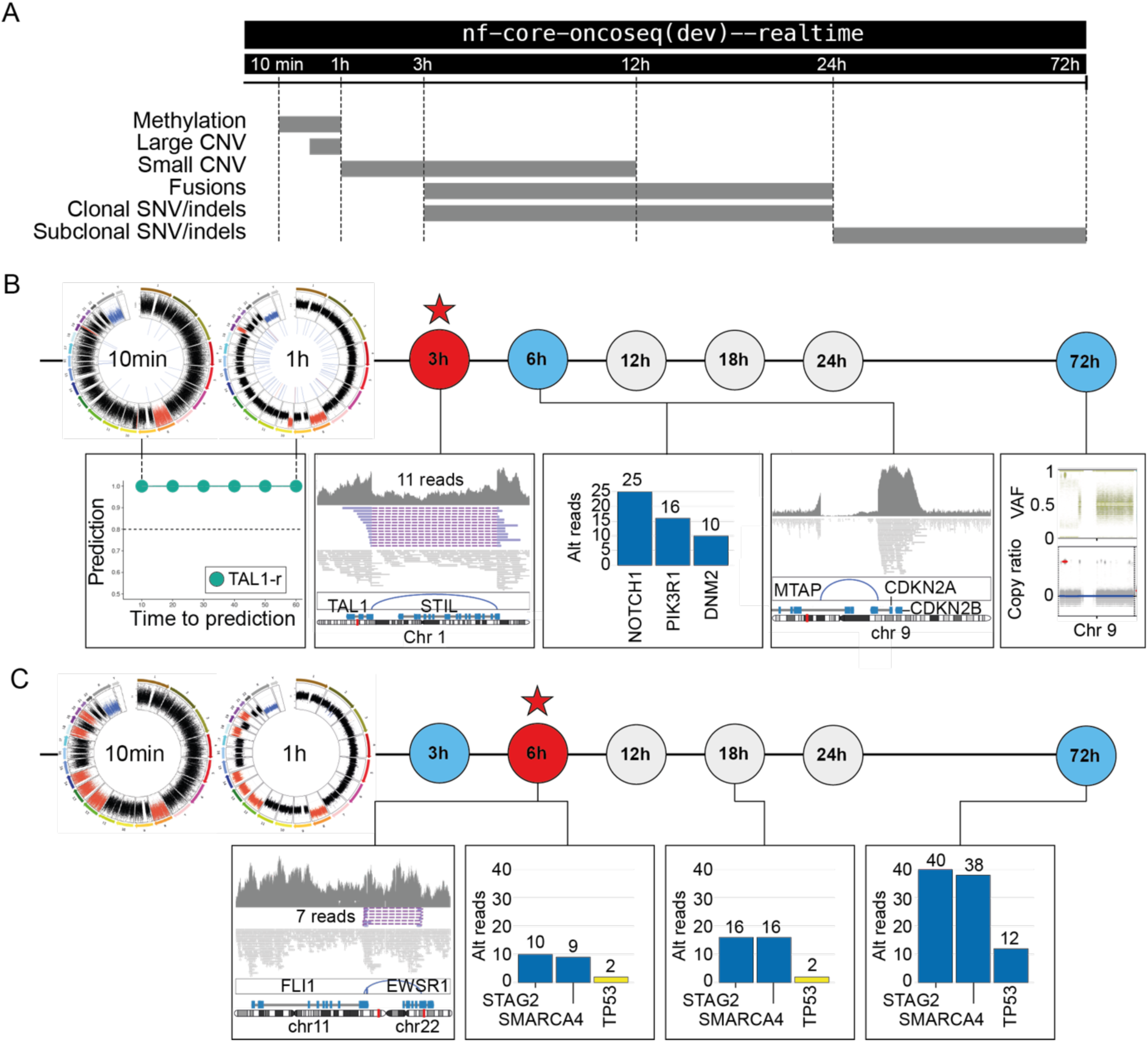
Timely classification and identification of alterations with workflow optimization for rapid precision diagnoses. **A)** Proposed timeline and sequencing order for optimal identification of all categories of alterations. **B)** Example 1: T-ALL sample (SI_ASWGS1009) showing a *STIL*::*TAL1* fusion, mutations in *NOTCH1*, *PIK3R1*, and *DNM2*, a *CDKN2A* deletion, trisomy of chromosome 8, gains of 10p and 19q, and CN-LOH on 9p. **C)** Example 2: Ewing sarcoma sample (SI_ASWGS 1258) with an *EWSR1*::*FLI1* fusion, clonal mutations in *STAG2* and *SMARCA4*, subclonal mutation in *TP53*, and trisomies of chromosomes 8, 12, 13, 14, 18, and 20. Note: The *TP53* mutation is not shown in Figure 2 due to a variant allele frequency (VAF) < 5% in the clinical WES (3%).

## DISCUSSION

Pediatric cancers are among the most genomically complex and heterogeneous diseases, driven by a wide spectrum of gene fusions, mutations, copy number alterations, and epigenetic changes. Intra-tumoral heterogeneity, in both clonal architecture and the tumor microenvironment, further complicates the detection of these alterations, particularly when tumor cell content is low, thereby necessitating higher-coverage approaches. Although conventional >80X short-read WGS remains the most comprehensive single assay for identifying clinically relevant genomic events, its relatively slow turnaround time prevents it from replacing other approaches (FISH, RT-PCR, etc.) that provide initial genomic insights. As a result, in practice, multiple sequential or parallel tests are still required to achieve timely and complete genomic profiling, which is material-intensive, resource-heavy, costly and not always feasible in urgent clinical contexts.

To address these limitations, we optimized and applied AS-WGS to provide a single, timely, and comprehensive assay capable of capturing the full spectrum of genomic alterations in pediatric cancers. While adaptive sampling has already demonstrated its utility in various biomedical and environmental contexts^13, 41–45^, its application in oncology is still relatively recent but most promising^9, 12^.

In our study, AS-WGS was uniformly applied on a cohort of 31 pediatric cancer patient samples, achieving a mean on-target coverage of ∼165X and off-target coverage of ∼18X, corresponding to an enrichment factor of ∼10X. Compared with extensive clinical testing, this depth enabled the reliable detection of key molecular and cytogenetic alterations across multiple tumor types, including leukemias and solid tumors. It allowed high-confidence identification of most alterations, including somatic variants down to variant allele frequencies >5%, as it is generally accepted for clinical NGS testing^46^.

Our results represent a substantial improvement over previously published AS-WGS datasets, advancing the development of a single comprehensive test for pediatric oncology. For example, Geyer *et al.*^12^ reported a target coverage of 86X in pediatric leukemia samples using panels of varying sizes. While sufficient for CNV and fusion detection, this coverage limited somatic mutation sensitivity to variants with VAF >30%, requiring a parallel NGS-based assay for mutation detection. Kato *et al.*^47^ achieved 21.3X coverage, with 40% recall of clinical SNVs and 18% recall of small indels. Other studies in germline and somatic cancer contexts also reported lower coverage and sensitivity, insufficient for confident detection of low-frequency variants^48–53^. In neuro-oncology, methylation profiling was used to categorize 50 samples in real-time, and AS-WGS on a 3487-gene panel provided additional diagnostic insights, including fusion and CNV detection, with ∼30X on-target coverage^15^. Taken together, our results are the first to achieve the sequencing depth needed to detect every type of genomic alterations, including mutations in pediatric cancers using a single assay. We also demonstrate that methylation-based classifiers, which provide valuable diagnostic information, are compatible with high coverage AS-WGS targeting gene panels for somatic alterations.

Our results provide valuable experimental insights into AS-WGS. We developed a protocol, based on ONT guidelines, using a ∼380-gene panel relevant to pediatric oncology. To our knowledge, this is the first AS-WGS cohort processed in a fully standardized manner, informing about important variables that affect performance. Data yield was partly dependent on flow cell performance, mainly linked to the number of active pores, and batch effects were observed when output was unexpectedly low. DNA quality also impacted results: samples with DIN >8 generally produced better data, but lower quality conditions can potentially be partially rescued: using long fragment buffer improved coverage (112X) despite a low DIN of 6.6 (Figure S4A). To reflect real-world clinical conditions, we included all consecutive samples in the analysis, even those with low DNA quantity or quality, which are often the only ones available. This comprehensive approach offers a realistic evaluation of AS-WGS performance across diverse sample conditions, with full metrics reported in Supplementary Table S4.

Among analysis pipelines developed for sequencing data, several, including the Illumina DRAGEN suite^54^ and nf-core-oncoanalyser^55^, are optimized for short-read data. The Epi2me suite of workflows^56^, although designed for nanopore sequencing, relies on internal standards that limit customization. Importantly, none of these pipelines supports time-resolved evaluation of adaptive sampling. As oncology-oriented tools tailored for tumor variant detection continue to mature, there is a growing need for flexible integration. To address this, we introduce a new workflow that supports adaptive sampling target handling and rapid onboarding of new tools (Figure S1), inspired by Epi2me’s architecture and aligned with nf-core standards^57^. Many tools used in this study require complete bioinformatics files as input. While Dorado and MinKNOW allow real-time streaming of some formats, they do not support live variant extraction. Retrospective analysis shows that clonal SNVs are detectable early, but current tools are too slow for real-time use. In contrast, methylation-based classification offers rapid insights, with molecular classes detectable within 10 minutes (Figure 4C). Although classifier results require orthogonal support to identify predicted drivers, this approach has strong potential for improvement as training datasets grow. Based on these findings, we developed a minimal version of our workflow to run selected tools at strategic timepoints, enabling rapid variant detection and flexible input handling depending on MinKNOW settings, while remaining lightweight enough to run directly on a variety of sequencing hardware.

A 72-hour AS-WGS experiment, including fragmentation and library reload, costs ∼1700 Canadian dollars and captures most of the information typically obtained through all cytogenetic and sequencing approaches that, combined, are significantly more expensive. A key advantage of AS-WGS is that all analyses are performed on a single platform. The compact size of the ONT sequencer also supports broader clinical adoption. Unlike probe-based approaches, AS-WGS allows same-day panel modifications, as targets are defined digitally, enabling case-by-case customization. Additionally, shallow background coverage preserves access to genome-wide SNP and methylation data.

Some limitations of AS-WGS were noted during our analysis. As AS-WGS is a PCR-free technique, it requires substantial starting material of ∼2 µg, and the need for fragmentation in adaptive sampling further increases this requirement. Background sequencing data consists of reads rejected after ∼700 bp, resulting in a significantly lower N50 compared to non-adaptive WGS (typically 15–20 kb), which may compromise phasing in low-complexity regions. Flow cell variability is another challenge: even among high-quality samples (DIN ≥9), target coverage varied widely (95X–251X), leading to variable genomic insights. Additionally, some fusions and SNVs were not detected by any software in the nf-core-oncoseq pipeline and required manual confirmation via genome browser visualization of split reads and point mutations. Complementary software tools should be evaluated and integrated to improve mutation detection.

## CONCLUSION

Our study demonstrates that high-coverage AS-WGS enables the detection of the full spectrum of clonal and subclonal alterations in pediatric cancers through a single, rapid and integrated assay. Real-time sequencing and adaptive analysis capabilities permit comprehensive molecular diagnoses to be achieved within the first hours of sequencing, a feature that will only become more powerful with ongoing technological and algorithmic advancements. It holds strong potential to positively impact the trajectory of pediatric cancer patients by enabling faster and more comprehensive, and accessible genomics-based clinical decision-making. While our approach was primarily developed for pediatric cancers, it is readily applicable, with same panel, to adult-onset malignancies that also affect the pediatric population, such as acute leukemias and sarcomas. Moreover, the target regions can be easily adapted to encompass genomic features relevant to other adult-onset cancer types. Therefore, this strategy holds significant potential for broader clinical application.

## Data Availability

All data produced in the present study are available upon reasonable request to the authors.

## AUTHOR CONTRIBUTIONS

Project design and development by VPL, with contributions from SC, THT, DS, MAS and AS. Bioinformatic development and analyses by CLD and NG. Data interpretation by SyL with contribution by CR. AS-WGS developed and performed by MAA. Additional bioinformatic analyses by ND, ACC, VL, BK, ARS, PTD and SF. Clinical genomics by AR, LJ, IB, VPL, SC and THT. Signature project coordination by ARB, and MPGI coordination by SeL. NG, CLD, SyL, VL, ND and VPL co-wrote the manuscript. Signature project co-leadership including patient recruitment: NJ, CG, SV, RS, SC, THT, VPL and DS. Sample coordination and biobanking by TS. All authors have read and approved the manuscript.

## RESEARCH SUPORT

The Signature project, including patient recruitment, biobanking, and data analysis, is funded by the Fondation Charles-Bruneau. Sequencing activities and additional experimental and bioinformatic staff support were made possible through the Azrieli Precision Child Health Platform. Funding was also provided by the Leukemia and Lymphoma Society of Canada. The MPGI also receives operational support from the Ministère de l’Économie de l’Innovation et de l’Énergie. Fondation Charles-Bruneau also support work from VPL, SC, THT, DS, MAS, NJ, CG, RS and SV. SC, VPL and MAS were supported by Génome Québec Integration Genomic Program. VPL, THT and RS are supported by Clinician-Scientist Scholarships from the Fonds de recherche du Québec – Santé. This research was enabled in part by support provided by Calcul Québec (www.calculquebec.ca) and the Digital Research Alliance of Canada (alliancecan.ca).

## ACKNOWLEDGEMENTS

The authors thank Sara Marullo, Manon Nayrac, and Roselle Gélinas for their contributions to project management of the Azrieli Precision Child Health Platform, as well as the CRA-CHUSJ biobank personnel under the supervision of Jocelyne Ayotte. Gratitude is also extended to Valérie Villeneuve, Chief of CRA-CHUSJ platforms, and to other members of the Integrated Genomics Meta-Platform team.

## Notes

### Competing Interest Statement

AS is a co-founder of NewCode Oncology, the developers of a cancer diagnostic platform licensed from the Hospital for Sick Children (SickKids).

### Author Declarations

Ethics committee of Centre Hospitalier Universitaire Sainte-Justine gave ethical approval for this work.

