## Supplementary Methods and Figures for "Single-workflow Nanopore whole genome sequencing with adaptive sampling for accelerated and comprehensive pediatric cancer profiling"

#### Cohort

Patients were enrolled in SIGNATURE cancer precision medicine program enrolling patients from all 4 pediatric oncology centers of the province of Quebec, Canada: Centre Hospitalier Universitaire Sainte-Justine, McGill University Health Centre, Centre Hospitalier Universitaire de Québec-Université Laval and Centre Hospitalier Universitaire de Sherbrooke. The institutional review board approved the research protocol, and written informed consent was obtained from all participants and their parents or legal guardians. Diagnostic tumor tissue was collected at the time of diagnosis through routine clinical procedures, including bone marrow aspiration, biopsy, or surgical resection.

#### Clinical genomics

**Whole exome-sequencing (WES).** Total DNA was extracted with the flexigene DNA kit (Qiagen) at the molecular diagnostics laboratory. WES was performed by the Centre Québécois de Génomique Clinique (CQGC). Libraries were generated from 100 ng of DNA using the KAPA HyperCap HyperPlus Workflow v.3.0 kit from Roche. The DNA probes used for capture (KAPA HyperExome) target approximately 43 Mb of the GRCh38/hg38 genome assembly. More specifically, they target both DNA strands corresponding to coding exon sequences based on the following sources: CCDS release 20, RefSeq (07-30-2018), Ensembl release 93, GENCODE release 28, and ClinVar (07-29-2018). Sequencing is performed on a NovaSeq 6000 in paired-end mode (2x100 bp) with a target depth of approximately 250X. Deconvolution, alignment, and variant identification were carried out using the software suite on the DRAGEN[1] (Illumina) server at the CQGC. Genetic variations present in the panel genes were analyzed by professionals at the Molecular Diagnostics Laboratory (SNV and Indel: exons and splice variants ±10 bp). Interpretation was based on the AMP-CAP-ASCO recommendations[2]. Evaluation of patient SI_ASWGS0808 went through research WES as systematic clinical exome sequencing was not yet implemented. For this patient genomic DNA was isolated using mini AllPrep DNA/RNA kits from Qiagen and exomes were captured using the Sure Select XT Clinical Research Exome V2 kit (Agilent) as per the manufacturer's instructions. Sequencing was performed at the CQGC with the same instrument and target depth as clinical WES. Bioinformatic analysis for research WES data was performed as described in previous work[3].

**RNA-sequencing.** Total RNA was extracted with the QIAamp RNA blood mini kit (Qiagen) at the molecular diagnostics laboratory. Libraries prepared from 200 ng of RNA using the Stranded Total RNA Ribo-Zero Plus kit (Illumina) were sequenced on the NovaSeq 6000 platform (Illumina) in paired-end mode (2x100 bp) by the Centre Québécois de Génomique Clinique (CQGC). Bioinformatics analysis, including deconvolution, alignment, and fusion identification, was performed using the DRAGEN[1] server at CQGC and its software suite (Illumina). The FusionCatcher[4] and Arriba[5] tools were also used when no fusion was identified. A minimum of 100 million fragments aligned to coding regions is required, along with a ribosomal RNA content of less than 30%. Interpretation of clinical genomics was based on the AMP-CAP-ASCO recommendations[2].

Sample Preparation

**DNA extraction**. Genomic DNA (gDNA) was isolated from tumor specimens using the AllPrep DNA/RNA Kit (Qiagen) at CHUSJ institutional biobank. gDNA with a DNA Integrity Number (DIN) greater than 8.5 was recommended as input material, with 2 µg undergoing fragmentation using a Covaris g-TUBE. Shearing was performed by centrifugation at 5800 rpm for 1 minute in 50 µl, following the manufacturer's instructions, to obtain ~12 kb fragments. Quality control was assessed using the Genomic DNA ScreenTape on the Agilent 4150 TapeStation system to verify fragment size, while quantification was performed with the Qubit dsDNA High Sensitivity Assay Kit.

**Library preparation and sequencing**. Library preparation was done following the manufacturer's protocol using the SQK-LSK114 kit from Oxford Nanopore Technologies (ONT) and Short Fragment Buffer. The final library, ranging from 60–500 fmol, was divided into two portions. Approximately 100–120 fmol was initially loaded onto a single FLO-PRO114M flow cell, and initial sequencing done for 40 hours. The flow cell was then washed and reloaded with the remaining library, allowing sequencing to continue for up to 72 hours total. Adaptive sampling was configured in enrichment mode to the target panels. Any deviations from the stated protocol as well as the specific panels used for each sample are indicated in Table S4.

Bioinformatics

**Bioinformatics Workflow.** An end-to-end Nextflow workflow was created using nf-core standards, nf-core-oncoseq[6] (main branch), for basecalling, mapping and variant calling (Figure S1A). To optimize variant calling in real time, tools were selected according to their speed and relevance at different timepoints. For all timepoints, basecalling and mapping were included if input was pod5 or fastq files, whereas mapping was skipped if input files were aligned BAM files. CNV calling, SV calling and differential coverage analysis were also included in all timepoints. For running when sequencing time is under 6h, methylation classification using MARLIN[7] was included. Between 6h and 72h of sequencing, SNP calling was added, and for the 72h timepoint, Subchrom[8] was added (Figure S1B). This reduced version of the workflow is available when using the branch dev of nf-core-oncoseq and using the parameter `--realtime TIMEPOINT`.

**Retrospective Time-Series Sampling.** To evaluate the detection of reads supporting a particular mutation, we took advantage of the read sequencing timestamps that are emitted by the Dorado[9] basecaller when using the BAM format. This information represents the sequencing start time for each read and is carried forward during alignment by minimap2[10].

For selected sequencing timepoints, 0-1h, 0-3h, 0-6h, 0-12h, and 0-18h, timestamps are extracted from the BAM files using Ontime[11]. For each timepoint, the same steps described in the Analysis section are carried out on the subsampled reads tagged with the selected timestamp. The read IDs that contribute to a particular variant are not always identified by the software in the output, which prohibited the exploration of emergent variant reads over time on a per read basis. To address this, we developed an analysis tool, VARCLOCK[12] that scans each individually mapped read (BAM) in a set of regions (BED) along a defined set of variants (VCF) to categorize each read as REFERENCE, VARIANT or OTHER (variants that are not within the list of alternate alleles reported at a particular site) and output timestamped, read-specific variant information. An auxiliary script was created to down-sample the reads covering the region-variant combination and tag them accordingly for manual inspection via e.g. IGV.

**Analysis.** All samples used in this study were processed in a standard way from POD5 files, using the nf-core-oncoseq workflow (main branch). Basecalling was performed using the Dorado[9] `sup@v5.0.0` model with `sup@v5.0.0_5mCG_5hmCG@v2` modifications. Mapping was done with methylation tags preserved using the option `'-y'` of Minimap2[10], using the soft masked GRCh38 Genome Reference from UCSC[13] on reads of average per-base Phred quality score of 10 or more. Single nucleotide variants and small indels were called with ClairS-TO[14] using the `ont_r10_dorado_sup_5khz_ssrs` model and Clair3[15] using the `r1041_e82_400bps_sup_v500` model and the `'--snp_min_af=0.05'` option. Large structural variants were called with Sniffles2[16] with the `'--minsupport-auto-mult 0.05'` and `'--phase'` options, along with the tandem repeat annotations BED file for the Hg38 reference for improved calling in repetitive regions. Resulting VCF files are annotated using SnpEff[17] with the GRCh38.p14 database and the hg38 Clinvar[18] database. Copy number variants are called with qDNAseq[19] and SubChrom[8] in both WGS and Panel modes, called directly on Clair3's output VCF files. Phasing of Clair3 and ClairS-TO annotated

VCF files is done with WhatsHap's[20] phase command. The aligned BAM file is then phased twice using both VCF files separately with WhatsHap's haplotag command, and haploblocks are determined for both resulting phased BAM files with WhatsHap's stats command. Structural variants are confirmed visually with Figeno[21]. When ground truth variants were not detected by the pipeline tools, manual inspection (via IGV) and additional programs were explored: DeepVariant[22] for SNV detection, Delly[23] for CNV and SV detection.

**Coverage calculation.** Mean coverage was calculated on target regions and on off target regions, the aligned BAM file is first split in two using the same BED file used in the adaptive sampling sequencing (including buffer region), with Samtools[24] view `'-b'` option. Mosdepth[25] with defaults parameters was used to calculate mean coverage on off-target regions using the BAM file resulting from the Samtools `'--unoutupt'` option. Mosdepth was then run three times with the `'-b'` option using the BED file containing the coordinates (excluding the 20kb buffer) on the BAM file resulting from the Samtools `'--output'` option, with different filters. First, the option `'-F 1540'` was used to include secondary alignments. Second, the option `'-F 1796'` was used to exclude secondary alignments, the default for Mosdepth. Lastly, the options `'-F 1796 -Q 60'` were used to calculate mean coverage when only including uniquely mapped reads. Mean coverage in figures and tables corresponds to the coverage calculated with default Mosdepth options.

### SUPPLEMENTARY FIGURES

Supplementary Figure S1

**nf-core-oncoseq workflow.** Schema summarizing the different tools and processes called within the nf-core-oncoseq workflow for the full version ran on all samples of the cohort (A), and for the "lite" version to run in realtime (B). Additional details: https://github.com/chusj-pigu/nf-core-oncoseq

#### Supplementary Figure 1

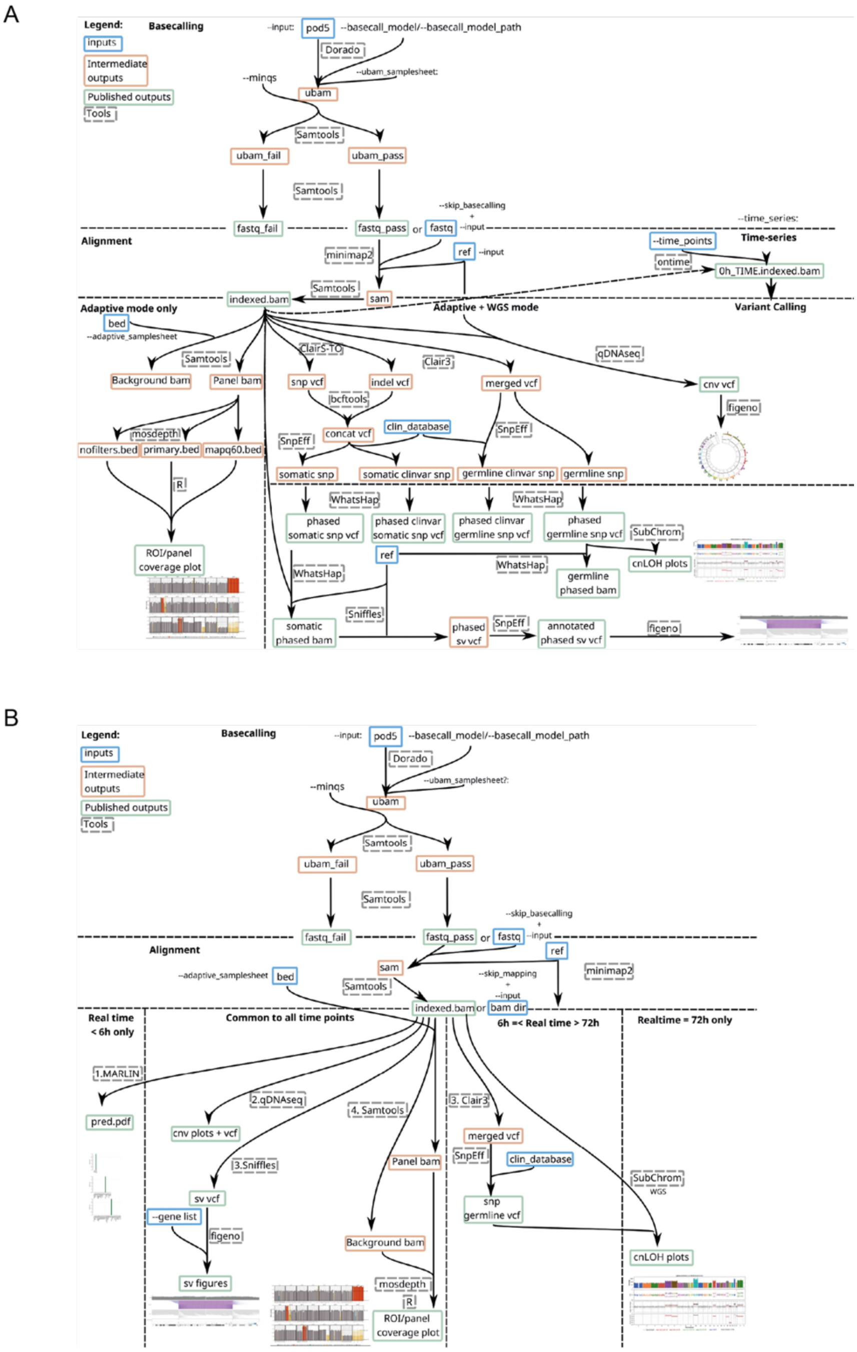

Supplementary Figure S2

**Coverage stability in copy neutral and non-copy neutral samples.** Gene level deletion and amplifications shown by differential coverage. Mean coverage of recurrently deleted genes *CDKN2A*/*B* and amplified gene *MYCN* shown across control and positive patients. Blue color represents a coverage with a Z score of < 2.75, red color represents a coverage with a Z score of > 2.75.

### Supplementary Figure 2

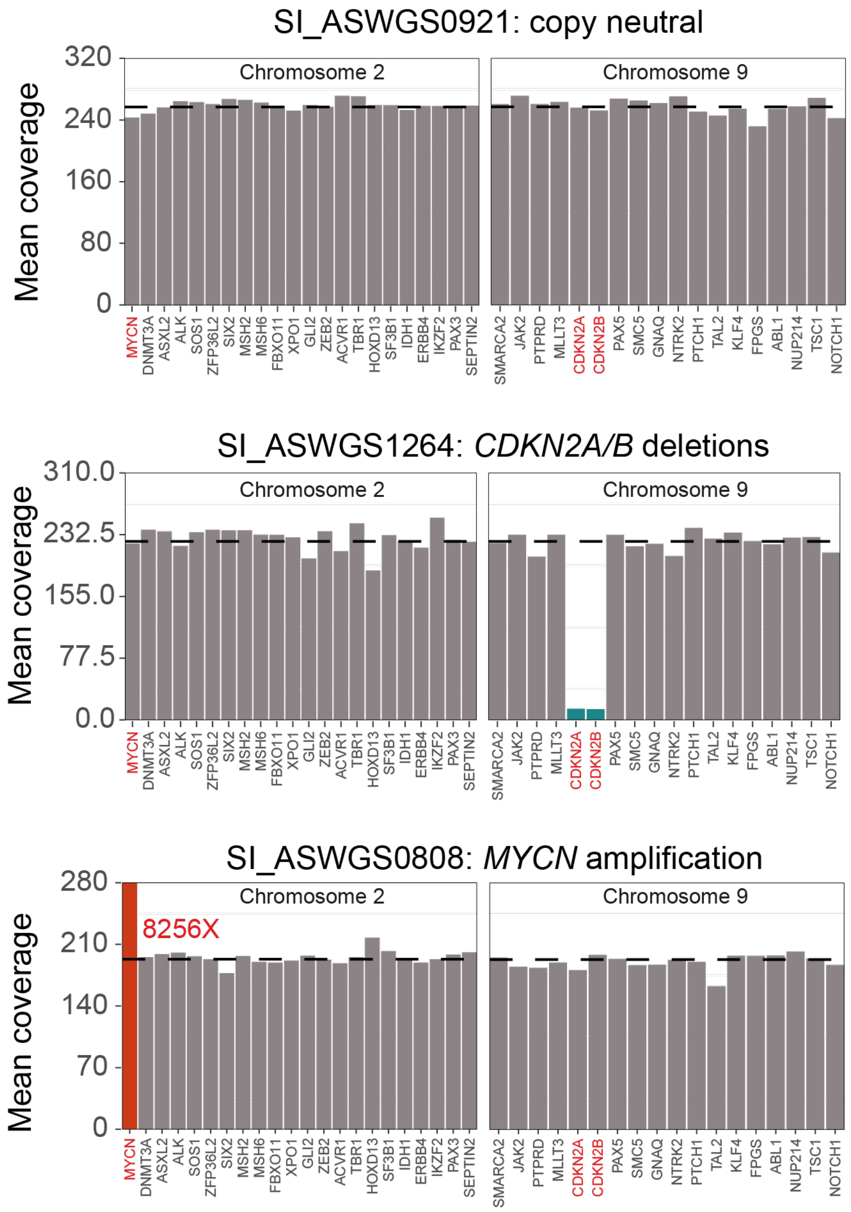

Supplementary Figure S3

**Mean coverage of on-target genes/loci.** Representative normal sample (SI_ASWGS0921 – mean coverage 186X). Genes are ordered by chromosomal position. Coverage of genes that exceed 1.5 times the median is capped to this value, and real value is annotated on the plot to facilitate visualization of variation. Alignment type filters are a combination of samtools flag filters and mapq filters. No filter is representative of all alignments including secondary alignments, primary only excludes secondary alignments but includes alignments of MAPQ= 0, and unique includes only alignments of MAPQ= 60. Dashed lines represent median on target (upper) and off target (lower) coverage.

### Supplementary Figure 3

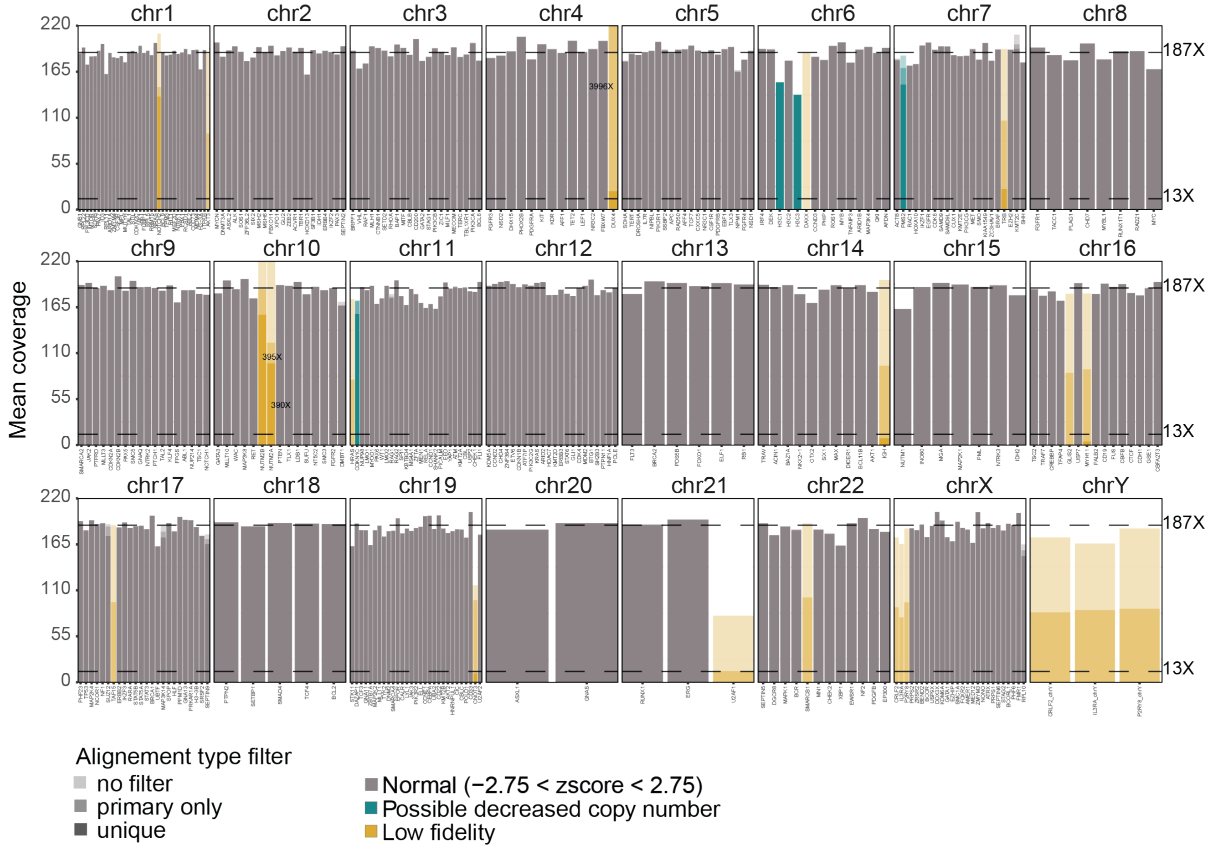

Supplementary Figure S4

**Coverage Statistics for the cohort. A**) Relationship between mean coverage in targeted region and gDNA DIN (n = 31), with point in grey representing the only sample for which a long fragment buffer was used to exclude short fragments (<3kb). **B**) Relationship between mean coverage inside of target (X) and number of flowcell pores at start of sequencing. Point color shows different dates of reception for the flowcell and point opacity showing input gDNA DIN. **C)** Inside of target mean coverage (X) across samples according to flowcell reception date to show variability between flow cell batches. Mann-Whitney test to compare mean coverage between dates of reception 24/07/2024 and 02/01/2025 resulted in a p value of 0.0003. Red dots identify the healthy donor sample that was sequenced twice.

**Supplementary Figure 4**

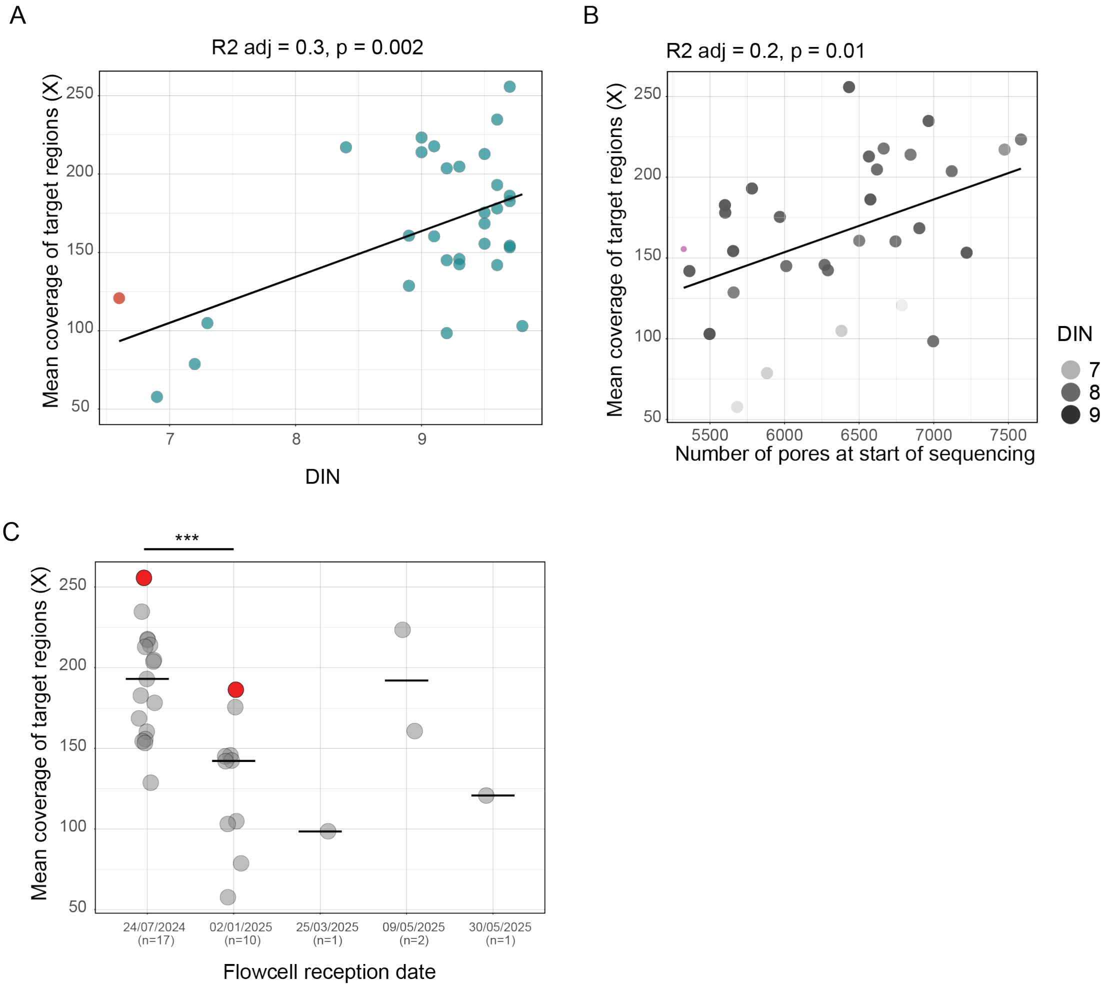

Supplementary Figure S5

Examples of visual confirmation of alterations missed by the workflow. **A)** *SET-NUP214* fusion in sample SI_ASWGS1002. **B)** *FLT3*-ITD (Internal tandem duplication) in sample SI_ASWGS1196. VAF WES: 11%; VAF AS-WGS: 7%).

### Supplementary Figure 5

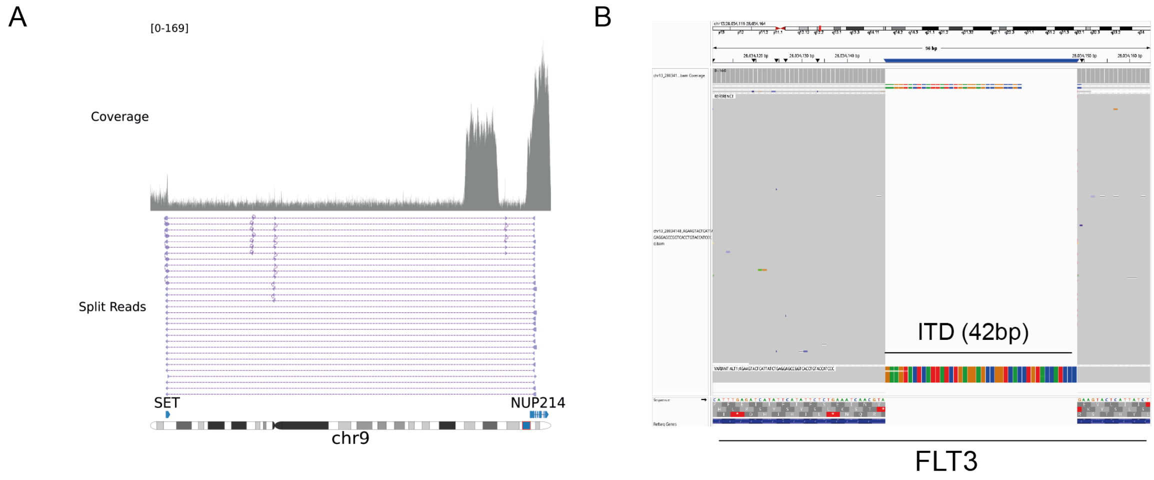

### SUPPLEMENTARY TABLES

Table S1

**Cohort description.** Comprehensive clinical description of the 30 patients undergoing AS-WGS.

Table S2

**Enrichment panel.** Genomic coordinates, gene/locus name and fidelity level for each selected region in the adaptive sampling panel.

Table S3

**On target and off target coverage from AS-WGS**. **A)** Mean coverage (X) inside each gene/locus in the adaptive sampling panel across each sample (n=31). Gene not included in the panel used for a specific sample is indicated with NA. **B)** Mean coverage (X) outside of the adaptive panel for each chromosome and across each sample (n=31).

Table S4

**Experimental details and quality control metrics.** Input DNA quality control metrics, flowcell characteristics and sequencing runs quality control metrics for each sample.

Table S5

**Genomic alterations of the cohort.** Mutational landscape of the cohort and comparison of AS-WGS findings with conventional diagnostic approaches. **A)** Single nucleotide variations (SNVs) and indels. **B)** Fusion. **C)** Copy number variations (CNVs) and loss of heterozygosity (LOH).

Table S6

**Methylation predictors.** ALMA and MARLIN prediction accuracy compared to clinical and molecular diagnosis.
